# Neurocognitive deficits, psychotrauma, and inflammation shape major depressive disorder and its phenome features

**DOI:** 10.64898/2026.02.11.26346056

**Authors:** Xueyan Wang, Ping Wang, Mengqi Niu, Chengkai Yangyang, Abbas F. Almulla, Chen Chen, Jing Li, Yingqian Zhang, Michael Maes

**Author notes:** Corresponding author: Prof. Michael Maes, M.D., Ph.D. International NIMETOX Center, Sichuan Provincial Center for Mental Health, Sichuan Provincial People’s Hospital, School of Medicine, University of Electronic Science and Technology of China, Chengdu 610072, China. https://scholar.google.co.th/citations?user=1wzMZ7UAAAAJ&hl=th&oi=ao Scholar GPS: Worldwide #1 in molecular neuroscience; #1/4 in pathophysiology Expert worldwide medical expertise ranking, Expertscape (December 2022), worldwide: #1 in CFS, #1 in oxidative stress, #1 in encephalomyelitis, #1 in nitrosative stress, #1 in nitrosation, #1 in tryptophan, #1 in aromatic amino acids, #1 in stress (physiological), #1 in neuroimmune; #2 in bacterial translocation; 3 in inflammation, #4-5: in depression, fatigue, and psychiatry.

## Abstract

**Background:** Major depressive disorder (MDD) involves immune–metabolic dysregulation, psychosocial adversity, and multidomain cognitive disturbances, yet single cognitive indices often show small and inconsistent effects. We derived a multivariate Cambridge Neuropsychological Test Automated Battery (CANTAB)-based cognitive phenotype (“cognitype”) and tested whether it adds explanatory value beyond adverse childhood experiences (ACEs) and an acute-phase protein (APP) index in acute-phase MDD.

**Methods:** Eighty-seven acute-phase MDD patients and 40 healthy controls completed CANTAB testing; key outcomes from DMS, RVP, OTS, and ERT were summarized as a cognitype score (PC1). ACEs were assessed, and peripheral inflammatory markers were combined into an APP index. Logistic and multiple regression models tested discrimination between MDD and controls and prediction of multidimensional phenome features (affective, physio-somatic, vegetative, recurrence-related, personality, and suicidality domains).

**Results:** Individual CANTAB outcomes showed limited between-group differences after FDR correction, but multivariable models integrating cognitive measures with ACEs and APP robustly discriminated MDD from controls (AUC up to 0.907). The cognitype independently predicted multiple phenome domains when modeled alongside ACEs and APP, and their combined effects explained ∼40–55% of variance across symptom dimensions.

**Conclusion:** A data-driven cognitype derived from core CANTAB tasks captures clinically meaningful cognitive variation in acute-phase MDD and contributes significant predictive value beyond psychosocial adversity and inflammatory activation. Integrating cognition, ACEs, and inflammation improves characterization of symptom heterogeneity and supports precision approaches targeting neurocognitive–immune–environmental mechanisms.

## INTRODUCTION

Major depressive disorder (MDD) is a highly prevalent psychiatric condition characterized not only by affective, vegetative and physio-somatic symptoms but also by marked disturbances in cognitive function. Cognitive impairment is increasingly viewed not as an epiphenomenon of mood disturbance but as a distinct cognitive phenotype of MDD, encompassing deficits in executive functioning, attention and emotion processing (McDermott & Ebmeier, 2009). These deficits persist beyond mood remission, contribute to poorer functional recovery, reduced occupational and social functioning, diminished quality of life, and an increased risk of subsequent neurodegenerative disorders (Jaeger et al., 2006; Majer et al., 2004). Importantly, cognitive impairments are present not only during acute episodes but also in first-episode patients and may persist into remission, indicating a trait-like component of neurocognitive vulnerability in MDD (Biringer et al., 2007; Conradi et al., 2011). However, cognitive findings in MDD remain heterogeneous and often show limited explanatory power, partly because many studies rely on isolated cognitive measures rather than integrated cognitive profiles. As a result, the nature and significance of an acute-phase MDD cognitive phenotype (i.e., a cognitype) remain incompletely understood.

A central unresolved issue is whether an MDD cognitype provides clinically actionable information about symptom expression. Recent work has argued that depression severity should be characterized using continuous, multidimensional symptom phenotypes rather than binary endpoints such as diagnosis status or single symptom totals. Maes et al. (2024) proposed modeling continuous indices capturing recurrence of illness, lifetime and current suicidal behaviors, and broader phenome features, which better reflect clinically meaningful heterogeneity. However, integrative models testing whether cognitive profiles predict these fine-grained symptom dimensions in acute-phase MDD remain scarce.

There is now evidence that MDD is a neuroimmune-metabolic-oxidative stress (NIMETOX) disorder (Maes et al., 2025). Nevertheless, it remains unclear how cognition relates to clinical symptom severity in a mechanistic sense. Growing evidence links positive (e.g., monomeric C-reactive-protein or mCRP) and negative (e.g., albumin and transferrin) acute phase inflammatory proteins, and oxidative markers to both MDD-related phenome and cognitive performance (Almulla et al., 2025; Bhattacharyya et al., 2025; Garés-Caballer et al., 2022; Maes, 1993; Maes et al., 2025). Immune-inflammatory and metabolic disturbances, such as pro-inflammatory cytokines and oxidative stress markers, can alter neural circuits supporting executive control and memory processes (Beckmann et al., 2022; Li et al., 2022), thereby contributing to cognitive deficits observed in depression (Wu & Zhang, 2023).

Beyond biological mechanisms, adverse childhood experiences (ACEs) represent a critical developmental risk factor for both depression, recurrence of illness, and NIMETOX pathways and long-term cognitive vulnerability (Maes et al., 2019; Zhang et al., 2024, 2023). Early-life trauma may induce long-lasting alterations in stress-related neurobiological systems and brain function, thereby increasing susceptibility to cognitive impairment and emotional dysregulation during depressive episodes (Xia et al., 2023).

Moreover, existing studies have provided important but fragmented evidence by examining inflammation, ACEs, and cognition largely in isolation. Both MDD and ACE exposure have been linked to impairments across distinct cognitive domains. Compared with healthy controls (HCs) and MDD patients without ACEs, individuals with MDD and ACEs exhibit elevated levels of inflammatory markers (Maes, 1995; O’Shields et al., 2025) and more pronounced cognitive deficits (Chakrabarty et al., 2020; Poletti et al., 2017). Moreover, a recent meta-analytic structural equation modelling study identified inflammation as a key mediator linking ACEs and MDD (Zagaria et al., 2024). Nevertheless, few studies have simultaneously modeled psychosocial adversity and immune–metabolic alterations as joint predictors of the acute-phase MDD cognitive phenotype, nor have they tested whether this cognitype contributes independently to clinical symptom burden.

In this sense, cognitive performance may operate as a downstream expression of psychosocial and biological vulnerability, potentially mediating the effects of ACEs and immune–metabolic dysregulation on symptom severity. Alternatively, the cognitype may capture a partially independent dimension of illness expression that predicts symptom burden beyond ACEs and biological indices. Distinguishing between these possibilities is essential for understanding whether cognitive phenotyping primarily indexes upstream vulnerability pathways or provides incremental clinical value as an independent predictor.

Building on this framework, the present study addressed two main questions in acute-phase MDD: (1) whether an empirically derived cognitive phenotype (cognitype), constructed from integrated cognitive performance measures, predicts the severity of multidimensional symptom outcomes (i.e., MDD phenome features, including recurrence of illness (ROI) and suicidal behaviors) beyond diagnostic classification; and (2) whether the cognitype mediates the predictive effects of ACEs and immune–metabolic dysregulation on symptom severity, or instead contributes independent explanatory power. To this end, we developed an integrative model linking psychosocial adversity (ACEs), immune–metabolic indices, cognitive performance, and continuous symptom phenotypes.

## METHODS

### Participants

This research is a cross-sectional case-control study. A total of 127 participants were enrolled, comprising 87 in the MDD study group and 40 in the healthy control group. Patients were enrolled in the Psychiatric Center of Sichuan Provincial People’s Hospital in Chengdu, China. The criteria for inclusion were as follows: (a) individuals aged 18-65 years of both genders; (b) written informed consent from all participants; (c) fulfillment of the diagnostic criteria for MDD as outlined in the Diagnostic and Statistical Manual of Mental Disorders, Fifth Edition (DSM-5); and (d) a Hamilton Depression Scale-21 (HAMD-21) score (Hamilton, 1960) exceeding 18. The healthy control group comprised staff members, family members of staff, and acquaintances of individuals with MDD. Controls were matched to patients by age, sex, education, and body mass index. The exclusion criteria for patients and controls included: (a) diagnoses of other significant mental disorders, such as bipolar disorder, schizophrenia, schizoaffective disorder, psycho-organic disorders, substance use disorders (excluding nicotine dependence), and autism spectrum disorders; (b) personality disorders (e.g., borderline, antisocial) and developmental disorders (e.g., severe intellectual disability), as well as neurological conditions like stroke, epilepsy, brain tumors, Parkinson’s disease,

Alzheimer’s disease, and multiple sclerosis; (c) major medical conditions, including autoimmune and immunological diseases, psoriasis, systemic lupus erythematosus, inflammatory bowel disease, rheumatoid arthritis, type 1 diabetes mellitus, chronic obstructive pulmonary disease, and cancer; (d) pregnant and lactating women; (e) individuals with a severe allergic reaction in the preceding month; (f) patients with a history of infection in the last three months; (g) patients undergoing treatment with immunosuppressive or immunomodulatory medications, including glucocorticoids; (h) individuals consuming therapeutic doses of antioxidants or omega-3 supplements within the past three months; (i) individuals with a surgical history in the last three months; or (j) frequent analgesic users. Moreover, healthy controls were excluded if they had a history of MDD or dysthymia (lifetime or current), any DSM-IV anxiety disorder, or a familial history of affective disorders, suicide, or substance use disorders (except nicotine dependency).

### Rating Scales

All participants were interviewed by a qualified researcher (physician) utilizing a semi-structured interview to evaluate demographic and clinical data, including age, gender, educational attainment, income, disease progression, medical history, frequency of depressive episodes, symptoms, personal history, and family history, among others. The Mini International Neuropsychiatric Interview (M.I.N.I.) (Sheehan et al., 1998) was utilized to screen for and diagnose major mental disorders and one personality disorder based on the criteria established in the DSM-IV and the International Statistical Classification of Mental Disorders (ICD-10). The scale is intended to assess both current and lifetime psychiatric disorders, which encompass: depressive episodes, dysthymia, hypomanic episodes, suicidal ideation, panic disorder, agoraphobia, social phobia (social anxiety disorder), generalized anxiety disorder, obsessive-compulsive disorder, post-traumatic stress disorder, alcohol dependence/abuse, non-alcoholic substance dependence/abuse, psychotic disorders, anorexia nervosa, bulimia nervosa, and antisocial personality disorder.

#### Clinical assessments

On the same day, the same evaluator administered scales to measure the severity of depression, anxiety, and physio-somatic symptoms. We utilized the total score from the 21-item HAMD scale to evaluate the severity of depression (Hamilton, 1960). We employed the Hamilton Anxiety Rating Scale (HAMA) scale score to assess the severity of anxiety (Hamilton, 1959). The Fibro-Fatigue Scale (FFS), a 12-item clinical interview, was utilized as a quantitative assessment of symptom intensity for Fibromyalgia and Chronic Fatigue Syndrome (CFS) (Zachrisson et al., 2002).

In addition to the clinician-related scales, participants completed self-rating scales to assess their depressive and anxiety symptoms, personality traits, and suicidal attempts and ideation. We used the Beck Depression Inventory (BDI-II) to measure self-reported depressive symptoms over the past 2 weeks (Beck et al., 1961), and the State-Trait Anxiety Inventory (STAI), state version, to evaluate self-reported anxiety (Spielberger, 2012), the Big Five Personality Inventory (John et al., 2012) to evaluate the personality in five trait domains, i.e., neuroticism, extraversion, openness, agreeableness, and conscientiousness. We focused on neuroticism, which reflects a stable tendency toward negative affectivity and heightened stress sensitivity. The Columbia-Suicide Severity Rating Scale (C-SSRS) (Posner et al., 2011) was employed to evaluate the cumulative instances of lifetime suicide attempts and suicidal ideation (Maes et al., 2024). We computed the recurrence of illness (ROI) index as a z-unit composite of (i) number of depressive episodes, (ii) lifetime suicide attempts, and (iii) lifetime suicidal ideation (up to one month before the index episode), derived from clinical interview and C-SSRS items as described previously (Maes et al., 2024).

Based on these clinical severity rating scales, we constructed integrated clinical severity indices as z unit-based composite or principal components scores reflecting the overall severity of depression (OSOD), affective symptoms, physio-somatic symptoms, vegetative symptoms, melancholia symptoms, current suicidal ideation (SI), neuroticism and recurrence of illness. We have previously explained why these clinical scores were selected and how they were computed (Niu et al., 2025). **Electronic Supplementary File (ESF), Table 1** summarizes briefly how these scores were computed.

**Table 1.** Socio-demographic and clinical data of patients with major depressive disorder (MDD) and healthy controls (HCs).

|  | HC (n = 40) |  | MDD (n = 87) |  | F/Chi's square | df | p-value |
| --- | --- | --- | --- | --- | --- | --- | --- |
|  | Mean | SD | Mean | SD |  |  |  |
| Sex (f/m) | 27/13 |  | 60/27 |  | 0.027 | 1, 127 | 0.869 |
| Age (years) | 37.080 | 13.749 | 34.070 | 11.600 | 1.634 | 1, 126 | 0.204 |
| Education (years) | 13.290 | 3.095 | 13.290 | 3.095 | 1126.000 | 1, 126 | 0.385 |
| Marital status | 19/17/1/2/1 |  | 36/43/1/7/0 |  | 3.366 | 1, 127 | 0.498 |
| Living conditions | 13/6/21/0 |  | 19/3/63/2 |  | 8.958 | 1, 127 | 0.030 |
| Work status | 27/1/3/0/9 |  | 35/30/5/1/16 |  | 16.485 | 1, 127 | 0.002 |
| Smoking | 37/3 |  | 71/16 |  | 2.555 | 1, 127 | 0.110 |
| MetS (Yes/No) | 29/10 |  | 72/14 |  | 1.516 | 1, 125 | 0.218 |
| MetS ranking | 1.590 | 1.464 | 1.407 | 1.231 | 0.524 | 1, 124 | 0.470 |
| BMI (km/m<sup>2</sup>) | 23.518 | 4.071 | 22.238 | 3.262 | 3.597 | 1, 126 | 0.060 |
| WC (cm) | 79.375 | 11.726 | 77.800 | 10.837 | 0.549 | 1, 126 | 0.460 |
| mCRP (mg/L) | -0.296 | 0.930 | 0.136 | 1.007 | 5.279 | 1, 126 | 0.023 |
| Albumin(g/L) | 43.641 | 1.986 | 41.120 | 3.190 | 20.668 | 1, 124 | <.001 |
| Transferrin (g/L) | 258.759 | 37.442 | 228.522 | 42.278 | 14.853 | 1, 125 | <.001 |
| APP index | -0.656 | 0.945 | 0.302 | 0.876 | 31.161 | 1, 126 | <.001 |
| ACEs | 30.975 | 9.189 | 42.955 | 12.520 | 29.304 | 1, 126 | <.001 |

All results are shown as mean (SD); MetS, metabolic syndrome; BMI, body mass index; WC, waist circumference; FF, Fibro-Fatigue; mCRP, high sensitivity monomeric C-reactive protein 9 assessed in logarithmic 34transformation; APP index, Acute phase proteins index; ACEs, adverse childhood experiences; Marital status, the number of participants who were single or unmarried/married/live together/widowed; Living conditions, number of participants who lived alone/with friends/with family (spouse, children, relatives)/others; Work status, the number of participants who were employed/unemployed/ retired/ housewife/full-time student.

#### Childhood trauma and metabolic syndrome (MetS)

We employed the Childhood Trauma Questionnaire-Short Form (CTQ-SF) to evaluate the severity of Adverse Childhood Experiences (ACEs) (Bernstein et al., 2003). Total ACE severity was computed as the sum of the five CTQ-SF subscale scores: emotional abuse, physical abuse, sexual abuse, emotional neglect, and physical neglect (Bernstein et al., 2003). This study utilized the aggregate of the five subscale scores to quantify the severity of all ACEs.

Body weight, height, heart rate, and blood pressure were evaluated. BMI was calculated as weight (kg)/height (m)². The waist circumference (WC) was measured in the horizontal plane, located halfway between the iliac crest and the lowest ribcage as an index of abdominal adiposity. MetS is characterized by the presence of three or more of the following components, as delineated in the 2009 Joint Scientific Statement issued by the American Heart Association and the National Heart, Lung, and Blood Institute: (a) A waist circumference of 90 cm or greater for males and 80 cm for females; (b) A triglyceride level of 150 mg/dL or higher; (c) A HDL cholesterol level of less than 40 mg/dL for males and less than 50 mg/dL for females; (d) Elevated blood pressure defined as greater than 130 mm Hg systolic or 85 mm Hg diastolic, or the use of antihypertensive medication; (e) An increase in fasting glucose levels of 100 mg/dL or more, or a diagnosis of diabetes (Alberti et al., 2009). Based on the number of MetS criteria, we used the MetS ranking in the analyses.

### Cognitive function assessments

Participants completed a CANTAB battery including One Touch Stockings of Cambridge (OTS), Rapid Visual Information Processing (RVP), Delayed Match to Sample (DMS), Match to Sample Visual Search (MTS), Spatial Working Memory (SWM), Emotional Bias Test (EBT), Emotional Recognition Test (ERT), and Cambridge Gambling Task (CGT). The 28 key outcome measures are summarized in ESF Table 2.

**Table 2.** Results of binary logistic regression with major depression (MDD) as the dependent variable.

|  | Explanatory Variables | B | S.E. | Wald (df=1) | Sig. | Exp(B) | 95% C.I. for EXP(B) |  |
| --- | --- | --- | --- | --- | --- | --- | --- | --- |
|  |  |  |  |  |  |  | Lower | Upper |
| Model #1 | DMS_PC0 | 0.888 | 0.255 | 12.12 | <.001 | 2.43 | 1.474 | 4.007 |
|  | ERT_UHRD | 1.065 | 0.284 | 14.091 | <.001 | 2.902 | 1.664 | 5.062 |
|  | OTS_MDLFC | 0.684 | 0.25 | 7.465 | 0.006 | 1.982 | 1.213 | 3.236 |
|  | RVP_MDL | 0.818 | 0.24 | 11.612 | <.001 | 2.265 | 1.415 | 3.626 |
|  | APP index | 1.546 | 0.287 | 28.952 | <.001 | 4.695 | 2.673 | 8.246 |
|  | ACEs | 1.234 | 0.236 | 27.445 | <.001 | 3.434 | 2.164 | 5.448 |
| Model #2<br>with age<br>& sex<br>entered | DMS_PC0 | 0.801 | 0.275 | 8.508 | 0.004 | 2.227 | 1.3 | 3.815 |
|  | ERT_UHRD | 0.65 | 0.311 | 4.368 | 0.037 | 1.916 | 1.041 | 3.524 |
|  | OTS_MDLFC | 1.072 | 0.328 | 10.684 | 0.001 | 2.921 | 1.536 | 5.555 |
|  | RVP_MDL | 1.186 | 0.293 | 16.331 | <.001 | 3.273 | 1.842 | 5.817 |
|  | APP index | 1.629 | 0.318 | 26.183 | <.001 | 5.099 | 2.732 | 9.515 |
|  | ACEs | 1.35 | 0.298 | 20.587 | <.001 | 3.859 | 2.153 | 6.915 |
|  | Sex | 0.281 | 0.56 | 0.252 | 0.616 | 1.325 | 0.442 | 3.974 |
|  | Age | -0.098 | 0.028 | 12.165 | <.001 | 0.906 | 0.858 | 0.958 |

DMS\_PC0, percent correct (0 second delay) of Delayed Match to Sample test; ERT\_UHRD, unbiased hit rate disgust of Emotion Recognition Task; OTS\_MDLFC, median latency to first choice of One Touch Stocking of Cambridge task; RVP\_MDL, median response latency of Rapid Visual Processing task; APP index, acute phase protein index; ACEs, adverse childhood experiences.

### Assays

#### Monomeric C-reactive protein (mCRP)

Serum mCRP concentration was measured with Enzyme-linked immunosorbent assay (ELISA) kits (BioVendor, Brno, Czech Republic). The sensitivity is 0.63 ng/mL, and the intra-assay and inter-assay coefficients of variation (CVs) are < 10% and < 15%, respectively.

#### Albumin

Serum albumin was measured using the Bromocresol Green Method kit (Beijing Strong Biotechnologies, Inc.) on a fully automated biochemical analyzer (ADVIA 2400, Siemens Healthcare Diagnostics Inc.), with intra- and inter-assay analytical CVs of 1.20% and 2.10%, respectively.

#### Transferrin

Serum transferrin was measured using an immunoturbidimetric assay (DIAYS DIAGNOSTIC SYSTEM (SHANGHAI) CO., LTD) on a fully automated biochemical analyzer (ADVIA 2400, Siemens Healthcare Diagnostics Inc) with a sensitivity of 0.03 g/L, intra-assay and inter-assay analytical (CVs) of 1.96% and 0.67% respectively.

### Statistics

Group differences in continuous variables were examined using analysis of variance (ANOVA). Where appropriate, p-values were adjusted using the false discovery rate (FDR). To discriminate MDD from healthy controls, we conducted binary logistic regression with diagnosis (MDD vs HC; HCs as reference) as the dependent variable, adjusting for age, sex, and years of education; these models used selected individual CANTAB outcomes as predictors. Model performance was evaluated using receiver operating characteristic (ROC) analysis (AUC with 95% CI). Multiple linear regression models were used to predict symptom and phenome scores from demographic/metabolic variables and biomarkers using two complementary strategies: (i) models including selected individual CANTAB outcomes, and (ii) parallel models replacing these outcomes with the cognitype score. Collinearity was assessed using tolerance and variance inflation factor (VIF). All tests were two-tailed with α = 0.05. Analyses were conducted in IBM SPSS Statistics (version 30.0). When needed, variables were normalized using log10, square root, rank, or winsorizing transformations.

## RESULTS

### Sociodemographic and clinical data

**Table 1** shows the sociodemographic and clinical data of the MDD patients and HCs in the current study. There was no significant difference in sex ratio, age, years of education, and smoking history between MDD and HCs. The MDD and HC groups either showed no significant differences in marital status, but did differ in living conditions and work status. There was no significant difference in metabolic variables, including MetS prevalence, MeaS ranking, BMI and waist circumference. Serum albumin and transferrin were significantly lower in the MDD patients, while mCRP and APP index were significantly higher than in HCs. The MDD patients showed considerably higher ACEs scores.

### CANTAB Features of MDD

ANOVA showed that only one CANTAB test result was (marginally) different between MDD and controls, namely OTS_MDLFC (controls: 13554±1275; MDD: 16145±865; F = 4.39, df = 1, 125, p = 0.038). However, after FDR p-correction, these differences were no longer significant. Nevertheless, after accounting for ACEs and the APP index in multivariable binary logistic regression analyses, significant associations between CANTAB tests and MDD emerged. **Table 2** shows two different logistic regression models predicting MDD as the dependent variable and HCs as the reference group. Model #1 indicates that MDD was significantly associated with APP index, ACEs and cognitive measures, including DMS_PC0, ERT_UHRD, OTS_MDLFC, and RVP_MDL (χ² = 127.787, df = 6, p < 0.001; Nagelkerke = 0.619). The accuracy of this model was 84.1% (sensitivity = 79.3% and specificity = 87.5%). Model #2, in which age and sex were entered as covariates, showed that MDD remained significantly associated with APP index, ACEs, and cognitive measures, including DMS_PC0, ERT_UHRD, OTS_MDLFC, and RVP_MDL (χ² = 142.734, df = 8, p < 0.001; Nagelkerke = 0.670). Thus, after adjusting for age and sex, all predictors in model #1 remained significant. Years of education were not significant in this model. The accuracy of Model #2 was 88.4% (sensitivity = 82.8%, specificity = 92.5%).

ESF Table 3 presents the results of multiple regression analysis with CANTAB test results in the logistic regression as dependent variables and demographic and clinical characteristics as explanatory variables. We first examined all four CANTAB test results that were significant in model #1 **Table 2** (binary logistic regression analysis) and secondary we examined whether any of the other CANTAB tests were associated with any of the predictors. In summary, ACEs were associated with the APP index, whereas associations between ACEs and individual CANTAB outcomes were not significant after FDR correction. The APP index impacted (marginally) one CANTAB test, namely ERT_UHRD, while metabolic variables (either MetS, MetS ranking, or WC) affected ERT_UHRD, RVP_MDL, MTS_FAMD, and CGT_DMQMT. These effects, however, were no longer significant after FDR p-correction.

**Table 3.**
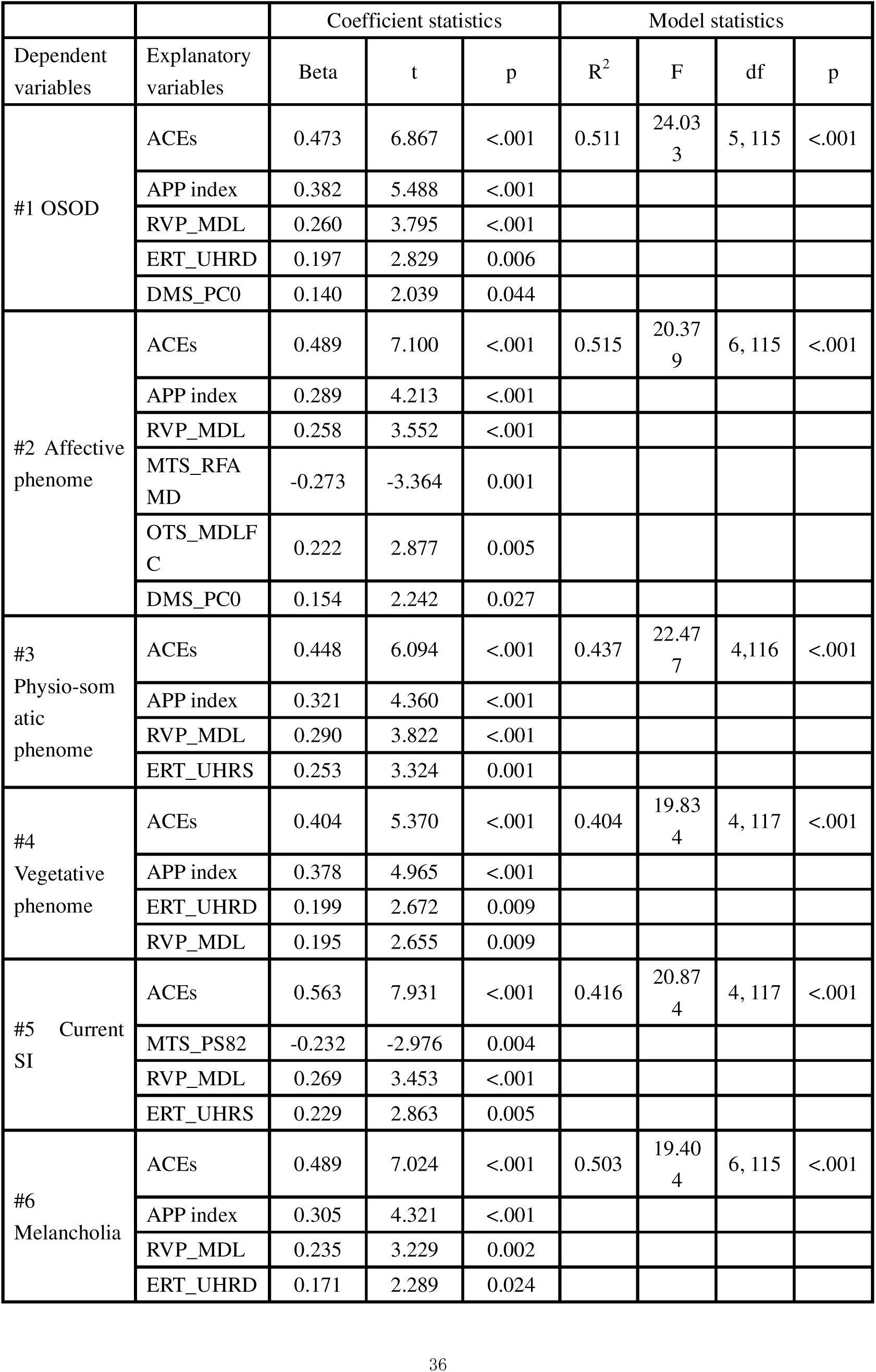
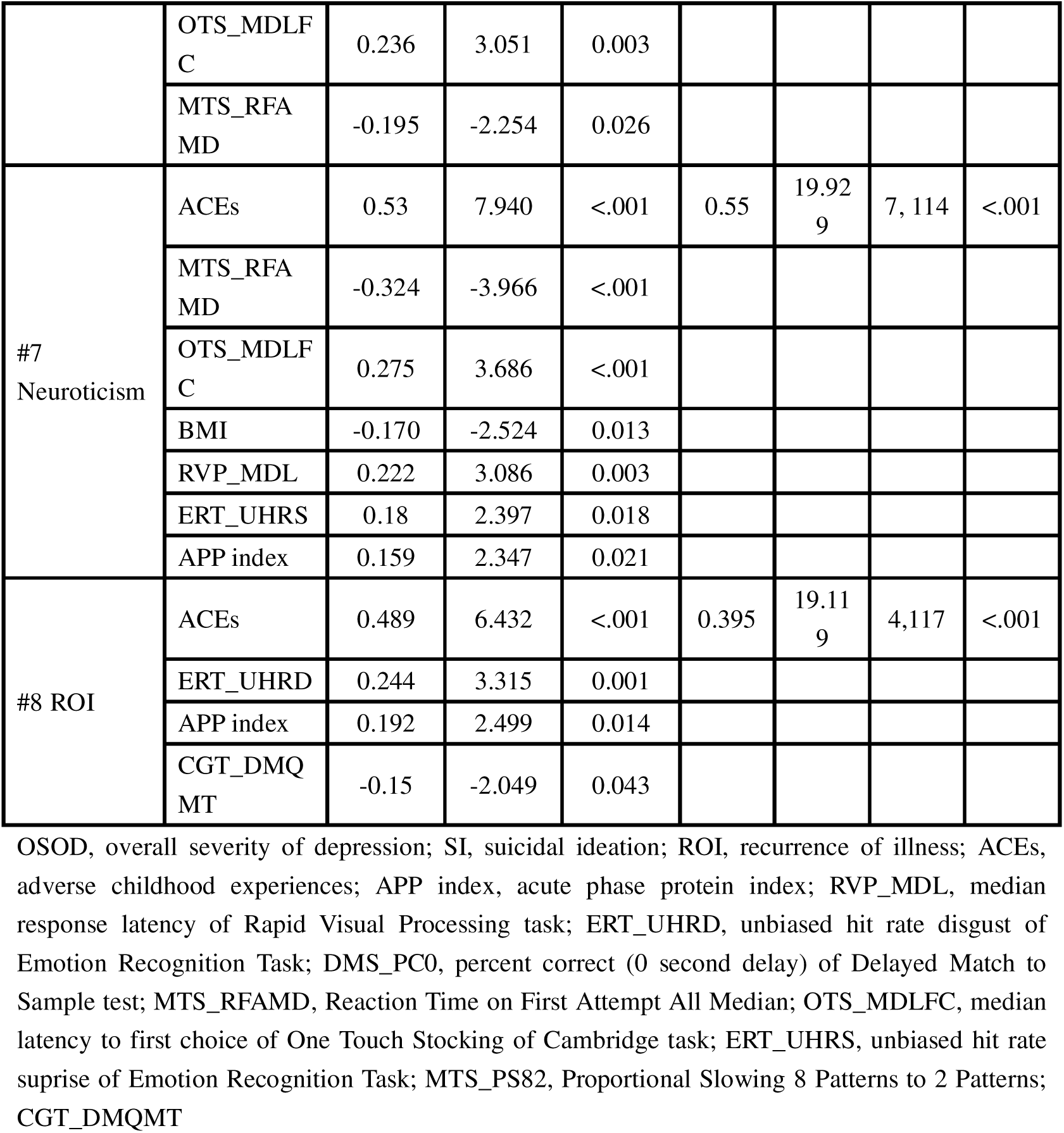
Results of multiple regression with the severity rating scales as dependent variables and neurocognitive, metabolic, and immunological biomarkers as explanatory variables.

### Prediction of the rating scale scores

**Table 3** shows the prediction of rating scale scores using neurocognitive, metabolic, and immunological data as explanatory variables. Model #1 shows that 51.1% of the variance in OSOD was explained by total ACEs, APP index, RVP_MDL, ERT_UHRD, and DMS_PC0 (all positive). Model #2 indicates that 51.5% of the variance in the affective phenome was explained by ACEs, APP index, RVP_MDL, OTS_MDLFC, and DMS_PC0 (all positive), while MTS_RFAMD contributed negatively. Model #3 shows that 43.7% of the variance in the physio-somatic phenome was explained by ACEs, APP index, RVP_MDL, and ERT_UHRS (all positive). Model #4 demonstrates that 40.4% of the variance in the vegetative phenome was explained by ACEs, APP index, ERT_UHRD, and RVP_MDL (all positive). Current suicidal ideation was predicted by ACEs, RVP_MDL, and ERT_UHRS (all positive), whereas MTS_PS82 contributed negatively, explaining 41.6% of the variance. Melancholia was predicted by ACEs, APP index, RVP_MDL, ERT_UHRD, and OTS_MDLFC (all positive), while MTS_RFAMD contributed negatively, together explaining 50.3% of the variance. Neuroticism was predicted by ACEs, OTS_MDLFC, RVP_MDL, ERT_UHRS, and APP index (all positive), while MTS_RFAMD and BMI contributed negatively, explaining 55.0% of the variance. ROI was predicted by ACEs, ERT_UHRD, and APP index (all positive), with CGT_DMQMT contributing negatively, explaining 39.5% of the variance.

**Table 4** summarizes multiple regression models predicting severity ratings from ACEs, the APP index, and the MDD cognitype. The latter was a highly significant predictor variable for all 8 clinical scores when combined with ACEs and the APP index (except for current SI). **Figure 1** shows the partial correlation between the z-unit composite socres of affective symptom and cognitype. **Figure 2** shows the partial correlation between the melancholia sympotom composite and the cognitype composite.

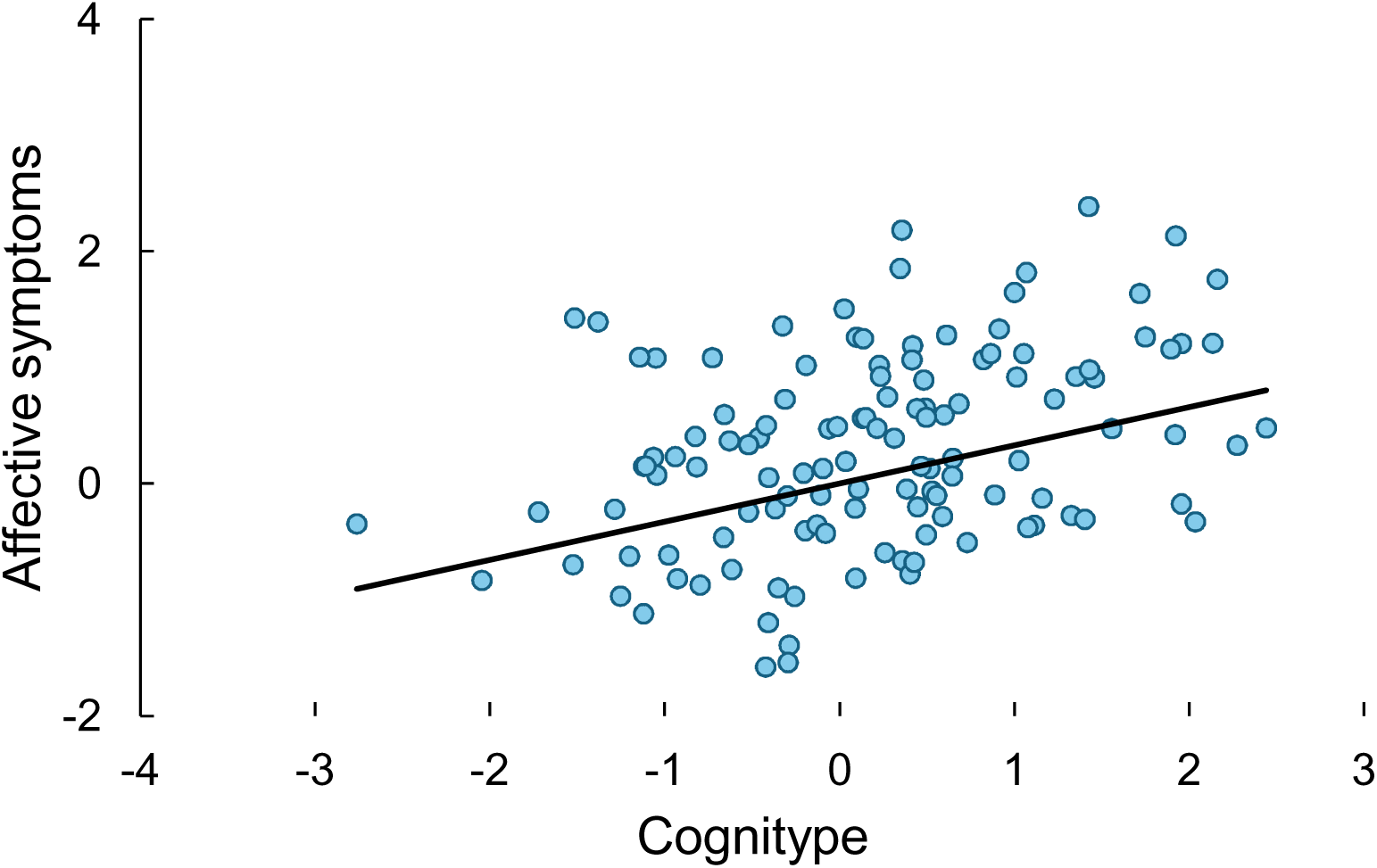

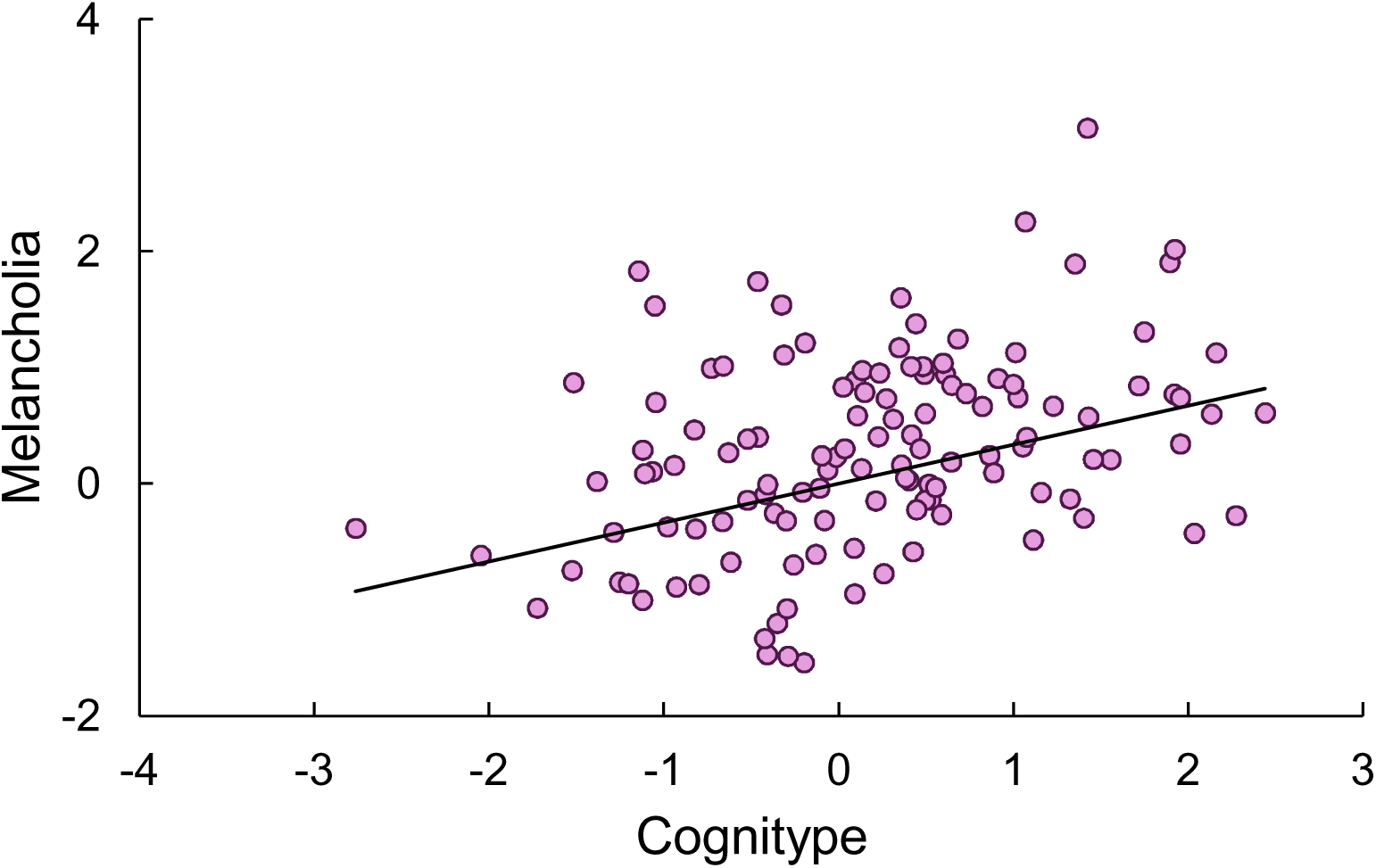

**Table 4.** Results of multiple regression with the severity rating scales as dependent variables and three explanatory variables, namely the cognitype, adverse childhood experiences (ACEs) and the acute phase protein (APP) index.

| Dependent variables | Explanatory variables | Coefficient statistics |  |  | Model statistics |  |  |  |
| --- | --- | --- | --- | --- | --- | --- | --- | --- |
|  |  | Beta | t | p | R<sup>2</sup> | F | df | p |
| #1 OSOD | ACEs | 0.446 | 6.66 | <.001 | 0.515 | 42.16 | 3, 119 | <.001 |
|  | APP index | 0.371 | 5.51 | <.001 |  |  |  |  |
|  | Cognotype | 0.332 | 5.17 | <.001 |  |  |  |  |
| #2 Affective phenome | ACEs | 0.469 | 6.81 | <.001 | 0.484 | 37.54 | 3, 120 | <.001 |
|  | APP index | 0.333 | 4.83 | <.001 |  |  |  |  |
|  | Cognotype | 0.291 | 4.41 | <.001 |  |  |  |  |
| #3 Physio-somatic phenome | ACEs | 0.418 | 6.094 | <.001 | 0.430 | 29.90 | 3, 119 | <.001<sup>e</sup> |
|  | APP index | 0.328 | 4.50 | <.001 |  |  |  |  |
|  | Cognotype | 0.300 | 4.31 | <.001 |  |  |  |  |
| #4 Vegetative phenome | ACEs | 0.378 | 5.26 | <.001 | 0.438 | 31.13 | 3, 120 | <.001<sup>e</sup> |
|  | APP index | 0.371 | 5.15 | <.001 |  |  |  |  |
|  | Cognotype | 0.317 | 4.61 | 0.009 |  |  |  |  |
| #5 Current SI | ACEs | 0.522 | 6.84 | <0.001 | 0.296 | 25.45 | 2, 121 | <.001<sup>e</sup> |
|  | Cognotype | 0.172 | 2.25 | 0.026 |  |  |  |  |
| #6 Melancholia | ACEs | 0.462 | 6.64 | <0.001 | 0.473 | 35.83 | 3, 120 | <.001<sup>g</sup> |
|  | APP index | 0.326 | 4.67 | <0.001 |  |  |  |  |
|  | Cognotype | 0.295 | 4.44 | 0.002 |  |  |  |  |
| #7 Neuroticism | ACEs | 0.531 | 7.37 | <0.001 | 0.435 | 30.82 | 3, 120 | <.001<sup>h</sup> |
|  | APP index | 0.221 | 3.06 | 0.003 |  |  |  |  |
|  | Cognotype | 0.219 | 3.18 | 0.002 |  |  |  |  |
| #8 ROI | ACEs | 0.483 | 6.31 | <.001 | 0.363 | 22.77 | 3, 120 | <.001<sup>e</sup> |
|  | APP index | 0.187 | 2.44 | 0.016 |  |  |  |  |
|  | Cognotype | 0.234 | 3.20 | 0.002 |  |  |  |  |

OSOD, overall severity of depression; SI, suicidal ideation; ROI, recurrence of illness; ACEs, adverse childhood experiences; APP index, acute phase protein index.

## DISCUSSION

This study addressed two central questions in acute-phase MDD: (i) whether an empirically derived neurocognitive phenotype (cognitype) captures clinically meaningful variance in symptom expression, and (ii) whether this cognitype may partly account for, or independently add to, the association of psychosocial adversity and immune–metabolic dysregulation with MDD symptom expression. Three main findings emerged. First, although univariate group contrasts of single CANTAB indices were modest after FDR correction, multivariable models integrating selected CANTAB outcomes with ACEs and the APP index robustly discriminated MDD from HCs even after demographic adjustment. Second, a composite cognitype derived from core CANTAB outcomes provided a compact cognitive summary and independently predicted multiple symptom phenomes when modeled alongside ACEs and APP. Third, integrated models combining cognition, ACEs, and APP explained substantial variance across phenome domains (≈40–55%), supporting a multidimensional framework for acute-phase MDD.

### Identification of an MDD cognitype

The cognitype was operationalized as the first principal component (PC1) extracted from four CANTAB outcomes (DMS_PC0, RVP_MDL, OTS_MDLFC, and ERT_UHRD). Previous studies of cognition in MDD have largely relied on single-task measures, leading to heterogeneous findings and limited explanatory power across samples and symptom domains (Gorlyn et al., 2006; Landrø et al., 2001; Murphy et al., 2003; Russo et al., 2015). By contrast, the present cognitype captures an integrated profile that more coherently represents acute-phase neurocognitive dysfunction. It was defined by three core features: (i) latency-based inefficiency in sustained attention and executive planning (higher RVP_MDL and OTS_MDLFC), (ii) relatively higher recognition accuracy at minimal delay (higher DMS_PC0), and (iii) heightened sensitivity to disgust-related emotional cues (higher ERT_UHRD).

The predominance of latency effects supports a profile of reduced processing efficiency in our severely depressed inpatients. Longer RVP_MDL is compatible with attentional fatigue and psychomotor slowing, whereas elevated OTS_MDLFC indicates slower planning and reduced cognitive-control efficiency. This pattern is consistent with meta-analytic evidence indicating that attentional and executive-control impairments represent core cognitive features of MDD (Bora et al., 2013; Kaser et al., 2017; McDermott & Ebmeier, 2009; Rock et al., 2014), and aligns with well-replicated deficits in attention (Luo et al., 2022; Schmidt et al., 2021; Weiland-Fiedler et al., 2004), executive function, and processing speed in MDD (Majer et al., 2004; Piani et al., 2022).

The relatively higher DMS_PC0 should not be interpreted as broadly “superior memory.” Within a multivariate configuration characterized by slowed control and attentional timing, preserved short-delay recognition accuracy may reflect compensatory recognition processes and/or altered response dynamics (e.g., conservative responding or a speed–accuracy trade-off) that coexist with reduced processing efficiency. This interpretation is consistent with models in which depression-related cognitive dysfunction primarily affects control and timing processes, while certain recognition components remain relatively intact under specific task demands.

Similarly, the elevated accuracy for disgust (ERT_UHRD) should not be interpreted as enhanced emotion recognition, which would contradict prior evidence of emotion-recognition deficits in MDD (Dalili et al., 2015; Mo et al., 2021). Instead, together with the meta-analytic evidence showing reduced accuracy for happy faces in MDD (Krause et al., 2021), higher disgust accuracy may reflect heightened sensitivity to negative, self-relevant cues. Disgust processing is clinically relevant because self-disgust and disgust-related negative self-referential processing are closely linked to shame, guilt, and self-criticism in depressive symptoms and affective disorders (Abdul-Hamid et al., 2014; Green et al., 2013; Rüsch et al., 2011). Consistent with negative-bias accounts of depression, elevated ERT_UHRD likely indexes increased emotional vigilance toward aversive stimuli rather than enhanced global emotion-processing capacity (Bourke et al., 2010).

Taken together, by integrating latency- and accuracy-based measures across cognitive and affective domains, the cognitype captures a system-level imbalance, executive and attentional slowing combined with heightened aversive salience and relatively preserved recognition accuracy, more effectively than isolated task analyses. This integrative approach helps reconcile inconsistencies in the single-task literature and supports cognitive phenotyping as a clinically meaningful dimension of acute-phase MDD.

### Neurocognitive-immune-metabolic predictors of the MDD phenome

Across affective, physio-somatic, vegetative, and personality-related phenomes, integrated models combining neurocognitive, immune, and psychosocial predictors explained a substantial proportion of variance (≈40–55%). ACEs emerged as a consistent predictor across all phenome domains, underscoring the broad and enduring impact of early adversity on emotional, somatic, and cognitive symptom expression in MDD. In parallel, the APP index and specific cognitive measures (e.g., RVP_MDL, DMS_PC0, ERT_UHRD) contributed differentially across symptom clusters, suggesting that distinct cognitive and biological mechanisms may underlie the heterogeneity of MDD expression. Together, this pattern supports a multidimensional view of MDD involving interacting neurocognitive, immune, and metabolic dysregulation rather than a purely affective disorder.

These results highlight cognitive phenotyping as a potentially practical tool for stratifying acute-phase MDD. The cognitype points to targets that are not identical to mood severity alone. For instance, latency-based executive inefficiency and negative-salience bias may inform personalized intervention selection (cognitive remediation, executive function training, emotion-bias modification) and may serve as mechanistically meaningful outcomes in trials. Furthermore, the strong combined prediction by ACEs + APP + cognitype motivates integrative assessment frameworks that treat cognition as a core clinical dimension rather than a secondary correlate.

### The cognitype independently predicts the MDD phenome

A central contribution of this study is that cognition provided predictive information not only for diagnosis but also for continuous symptom phenotypes. Using complementary approaches, models retaining individual CANTAB outcomes and models substituting these outcomes with the cognitype composite, we found that the cognitype accounted for significant unique variance in several phenome domains after ACEs and the APP index were included. In the parsimonious cognitype + ACEs + APP models, explained variance was substantial for OSOD (R² = 0.515), affective symptoms (R² = 0.484), physio-somatic symptoms (R² = 0.430), vegetative symptoms (R² = 0.438), melancholia (R² = 0.473), neuroticism (R² = 0.435), and ROI (R² = 0.363). For current suicidal ideation, ACEs and cognitype remained significant predictors, whereas the APP index did not contribute additional explanatory power (R² = 0.296).

A priori possibility was that cognition might primarily reflect a downstream pathway through which psychosocial adversity and immune–metabolic dysregulation shape acute-phase symptom expression. This hypothesis was supported by evidence linking ACEs to long-term stress sensitization and cognitive–emotional vulnerability (Xia et al., 2023; Zhang et al., 2024, 2023), and by studies implicating immune-inflammatory and metabolic disturbances in executive and memory-related processes (Beckmann et al., 2022; Bhattacharyya et al., 2025; Garés-Caballer et al., 2022; Li et al., 2022; Wu & Zhang, 2023). In addition, inflammation has been identified as a key mediator between ACEs and MDD severity (Zagaria et al., 2024).

However, across fully adjusted models, cognitype indicators remained significant predictors of multiple phenome domains after accounting for ACEs and the APP index, arguing against a mediator-only account. Instead, these findings support cognition as a partially independent dimension contributing to acute-phase MDD heterogeneity. When symptom expression is modeled as a continuous, multidimensional phenome, cognitive phenotyping provides explanatory information beyond mood state alone. Consistent with phenome-based frameworks (Maes et al., 2024), incorporating cognitype alongside psychosocial and immune–metabolic indices improves characterization of acute-phase symptom heterogeneity and yields information relevant to clinically salient outcomes such as melancholia and recurrence.

### Executive slowing, affectively hyper-responsive salience and brain circuits

Building on the cognitype features described above, the pattern observed here is more consistent with reduced processing efficiency than global cognitive decline, combining executive/attentional slowing with relatively preserved short-delay recognition accuracy and heightened aversive salience. At the circuit level, prolonged RVP latency plausibly reflects reduced efficiency within distributed executive-control systems that integrate prefrontal and cingulate control processes with parietal and temporo-associative regions and altered coupling with default-mode and salience networks implicated in depression (Pan et al., 2020; Piani et al., 2022). Increased OTS latency similarly points to slowed planning and reduced cognitive-control efficiency supported by fronto-cingulate circuitry, particularly dorsolateral prefrontal cortex (DLPFC) and anterior cingulate cortex (ACC); in this context, the co-occurrence of OTS slowing with preserved DMS accuracy is more compatible with executive inefficiency than with a primary perceptual deficit (Causse et al., 2019). Higher DMS_PC0 may reflect preserved recognition processes and/or adaptive response strategies under minimal delay, potentially relying on occipito-temporal and medial temporal pathways and hippocampal–prefrontal interactions. Elevated disgust recognition is closely tied to insula-centered salience circuitry with contributions from orbitofrontal and amygdala–prefrontal pathways, consistent with enhanced allocation of salience to aversive/self-relevant cues in depression.

This configuration provides a plausible bridge to symptom expression. Heightened sensitivity to disgust-related cues aligns with evidence for reduced processing of positive facial expressions and a relative bias toward negative affective information in MDD (Krause et al., 2021). Concurrent fronto-striatal inefficiency may delay reappraisal and weaken adaptive emotion regulation (Furman et al., 2011; Kang et al., 2016), thereby reinforcing negative bias and rumination and contributing to blunted reward responsivity (Harmer et al., 2009; Nakamura et al., 2018). The same fronto-striatal inefficiency profile is consistent with psychomotor slowing and fatigue-related phenotypes (Buyukdura et al., 2011), while reward-circuit hypoactivity (ventral striatal and orbitofrontal imbalance) may further reduce energy, desire, and appetite (Ng et al., 2019). In addition, altered integration of affect and nociception within amygdala–PFC control nodes may amplify pain perception, contributing to physio-somatic complaints (Zheng et al., 2022). This framework also accommodates preserved recognition accuracy alongside reduced throughput: motivational and effort-valuation alterations can reduce willingness to exert effort despite intact capacity (Treadway et al., 2012), and prefrontal inefficiency can increase perceived cognitive effort, intensifying fatigue during executive demands (Wagner et al., 2006).

The cognitype and related circuit-level alteration may further inform suicidal and personality-related symptom dimensions. Fronto-striatal slowness (higher OTS_MDLFC and RVP_MDL) implies delayed decision processes and compromised cognitive control, consistent with despondency and indecision in suicidal individuals (Yun et al., 2023).

Hyperreactive limbic responses and ventromedial prefrontal dysfunction can heighten negative emotional salience and perceived distress, strengthening self-referential rumination (Nakamura et al., 2018), while reduced dorsolateral prefrontal modulation weakens top-down inhibition and disrupts value-based decision-making (Furman et al., 2011). In parallel, neuroticism converges on similar circuitry: sustained medial prefrontal responses to sad faces suggest persistent negative self-referential processing (Haas et al., 2008), acute stress can amplify the impact of neuroticism on amygdala threat responsivity (Everaerd et al., 2015) and converging lesion/neuroimaging evidence links higher neuroticism to reduced DLPFC efficiency and weaker top-down control (Forbes et al., 2014), a pattern compatible with prolonged deliberation latency and heightened negative appraisal. Together, these findings support a model in which the cognitype links circuit-level inefficiency and affective hyper-responsiveness to both behavioral tendencies and clinically salient symptom dimensions in acute-phase MDD.

### Limitations and future directions

Several limitations should be acknowledged. First, the cross-sectional design precludes causal inference regarding the temporal relationships among psychosocial adversity, immune–metabolic activation, cognitive efficiency, and symptom phenotypes. Although the sample size was sufficient for the present multivariate analyses, replication in longitudinal cohorts will be important to confirm the stability and generalizability of the identified cognitype and its predictive contributions across clinical subgroups and analytic approaches.

Integrating structural and functional MRI with cognitive and immune–psychosocial variables could further enhance knowledge on the distinct brain circuits involved. Accordingly, the neural interpretations offered here are grounded in prior literature and should be considered hypothesis-generating. Subsequent multimodal analyses will build on the present cognitive framework to directly test the neural implementation of latency-based inefficiency and affective salience using structural and functional MRI.

## CONCLUSION

In conclusion, this study delineates a data-driven acute-phase cognitive phenotype of major depressive disorder characterized by latency-based inefficiency in executive and attentional processing, relatively preserved recognition accuracy under minimal delay and heightened sensitivity to disgust-related emotional cues. Although individual cognitive measures showed limited discriminatory power in univariate analyses, their integration with ACEs and the APP response robustly differentiated MDD from healthy controls. Beyond diagnostic classification, cognitive performance—summarized either by selected CANTAB outcomes or by a parsimonious composite cognitype—explained substantial and clinically meaningful variance across multidimensional symptom phenomes, including affective, physio-somatic, vegetative, personality-related features, melancholia, and recurrence of illness. Our findings support cognition as an independent dimension of acute-phase MDD heterogeneity rather than merely a downstream mediator. Together, these findings reinforce a biopsychosocial framework in which environmental risk, inflammatory processes, and neurocognitive dysfunction contribute through partially distinct yet converging pathways to shape the acute-phase expression of MDD.

## Ethical approval and consent to participate

This study obtained approval from the ethics committee of Sichuan Provincial People’s Hospital “Ethics (Research) 2024-203” and was executed in strict compliance with ethical standards and privacy regulations. All participants provided informed consent.

## Author’s contribution

XW performed the research, analyzed the data, and wrote the original draft and subsequent revisions. PW and AA contributed to writing—review and editing. MN coordinated the project. YY and JL recruited participants. CC performed the research. MM designed and supervised the study, secured funding, analyzed the data, and contributed to writing—review and editing.

## Availability of data

The data supporting the findings of this study can be obtained upon request from the corresponding author. The data are inaccessible to the public owing to privacy or ethical constraints.

## Supporting information

Electronic Supplementary File

