## Supplementary material for "Neurocognitive deficits, psychotrauma, and inflammation shape major depressive disorder and its phenome features": Electronic Supplementary File

Supplementary tables

Supplenmentary Table 1. Constructin of the clinical scores.

| **Items** | **Computation** |
| --- | --- |
| OSOD | Overall severity of depression, constructed as z score of a principal component from vegetative, affective, and physiosomatic symptom domains. |
| Affective phenome | A PC extracted from pure HAMD, pure HAMA, STAI-state, and pure BDI. |
| Physio-somatic phenome | A PC extracted from three somatic symptom domains which shape the single group factor in Table 1, namely SSS-8, pure CFS and pure physiosomatic. |
| Vegetative phenome | Sum of anorexia, early awakening, mental retardation, loss of weight, and diurnal variation (Hamilton Depression Scale). |
| Current SI | A PC extracted from six items of C-SSRS assessing current suicidal ideation intensity and frequency. |
| Melancholia | Z composite score: sum of z scores from HAMD (Q6 + Q8 + Q16 + Q17 + Q18) and BDI (Q4 + Q5 + Q6 + Q7 + Q8 + Q10 + Q12 + Q13 + Q18 + Q20). |
| Neuroticism | Sum of self-report items indexing emotional instability, anxiety, and negative affect (Big Five Inventory), with reverse-coded items recoded. |
| ROI | Z composite score: z (number of depressive episodes) + z (frequency of suicidal ideation) + z (frequency of suicidal attempts). |

OSOD, overall severity of depression; SI, suicidal ideation; ROI, recurrence of illness.

Supplenmentary Table 2. Key measures of CANTAB assessments.

| Task | Measure Name | Measure Description | Description | Sense | Units | Variant Applicability | Min | Max |
| --- | --- | --- | --- | --- | --- | --- | --- | --- |
| CGT | CGTDMQMT | Decisin Making Quality Total Merged | The proportion (0 – 1) of all trials where the subject chose the majority box color. Calculated over all assessed trials from both the ascending and descending conditions in which the number of boxes of each color differed. | +ve | - | CGT Ascending/Descending First | 0 | 1 |
| CGT | CGTRAJMT | Risk Adjustment Merged | Risk adjustment is a measure of sensitivity to risk, based on the ability to modify choices in the light of information about the probability of different outcomes and to track the optimal outcome on each trial. The measure is calculated from the average proportion of points that the subject chose to bet with, taking into account the number of colored boxes in the majority. | cx | - | CGT Ascending/Descending First | -5.4 | 5.4 |
| CGT | CGTDAVT | Delay Avension Total | Allows for the dissociation between risk taking and impulsivity by determining whether subjects simply just place a bet at the first opportunity. Calculated as CGT Risk Taking for all trials from the descending condition minus CGT Risk Taking for all trials from the ascending condition. | cx | - | CGT Ascending/Descending First | -0.9 | 0.9 |
| DMS | DMSPCAD | Percent Correct (All delays) | The percentage of assessment trials containing a delay during which the subject chose the correct box on their first box choice. Calculated across all assessed trials contraining a delay. | +ve | % | Recommended Standard | 0 | 100 |
| DMS | DMSPCS | Percent Correct (Simultaneous) | The percentage of assessment trials where the target and response stimuli were presented simultaneously during which the subject chose the correct box on their first box choice. Calculated across all assessed trials containing the simultaneous presentation of stimuli. | +ve | % | Recommended Standard | 0 | 100 |
| DMS | **DMSPC0** | Percent Correct (0 Second Delay) | The percentage of assessment trials containing a zero second delay during which the subject chose the correct box on their first box choice. Calculated across all assessed trials containing a zero second delay. | +ve | % | Recommended Standard | 0 | 100 |
| DMS | DMSPC4 | Percent Correct (4 Second Delay) | The percentage of assessment trials containing a four second delay during which the subject chose the correct box on their first box choice. Calculated across all assessed trials containing a four second delay. | +ve | % | Recommended Standard | 0 | 100 |
| DMS | DMSPC12 | Percent Correct (12 Second Delay) | The percentage of assessment trials containing a twelve second delay during which the subject chose the correct box on their first box choice. Calculated across all assessed trials containing a twelve second delay. | +ve | % | Recommended Standard | 0 | 100 |
| EBT | EBTBP | Bias point | The proportion of assessed trials where the subject selected 'Happy', adjusted to a scale of 0 to 15. (Number of assessed trials selected as 'Happy'/Number of all assessed trials)*15 | Higher scores indicate a bias towards chossing "Happy" | - | 1 | 0 | 15 |
| ERT | ERTRT | Median Reaction Time | The median latency for a subject to select an emotion after being presented with a stimulus. Calculated across all assessed trials. | Higher is Better | ms | 1 | 0 | ∞ |
| ERT | ERTTH | Total Hits | The total number of times the subject selected the correct emotion. Calculated across all assessed trials. | Higher is Better | - | 1 | 0 | 90 |
| ERT | ERTUHRA | Unbiased Hit Rate Anger | The unbiased hit rate ensures that recognition accuracy of the Anger emotion is not influenced by response guessing or response bias effects. It takes into consideration the joint probability of an individual making a correct response, based on the presentation of the correct stimulus out of the available possibilities. Calculated for assessed Anger trials only. | Higher is Better | - | 1 | 0 | 1 |
| ERT | **ERTUHRD** | Unbiased Hit Rate Disgust | The unbiased hit rate ensures that recognition accuracy of the Disgust emotion is not influenced by response guessing or response bias effects. It takes into consideration the joint probability of an individual making a correct response, based on the presentation of the correct stimulus out of the available possibilities. Calculated for assessed Disgust trials only. | Higher is Better | - | 1 | 0 | 1 |
| ERT | ERTUHRF | Unbiased Hit Rate Fear | The unbiased hit rate ensures that recognition accuracy of the Fear emotion is not influenced by response guessing or response bias effects. It takes into consideration the joint probability of an individual making a correct response, based on the presentation of the correct stimulus out of the available possibilities. Calculated for assessed Fear trials only. | Higher is Better | - | 1 | 0 | 1 |
| ERT | ERTUHRS | Unbiased Hit Rate Sadness | The unbiased hit rate ensures that recognition accuracy of the Sadness emotion is not influenced by response guessing or response bias effects. It takes into consideration the joint probability of an individual making a correct response, based on the presentation of the correct stimulus out of the available possibilities. Calculated for assessed Sadness trials only. | Higher is Better | - | 1 | 0 | 1 |
| ERT | ERTUHRSU | Unbiased Hit Rate Surprise | The unbiased hit rate ensures that recognition accuracy of the Surprise emotion is not influenced by response guessing or response bias effects. It takes into consideration the joint probability of an individual making a correct response, based on the presentation of the correct stimulus out of the available possibilities. Calculated for assessed Surprise trials only. | Higher is Better | - | 1 | 0 | 1 |
| MTS | MTSCFAPC | Correct Trials on First Attempt All Percentage | The percentage of trials during which the subject selected the correct box. Calculated across all assessed trials. | +ve | % |  | 0 | 100 |
| MTS | MTSPS82 | Proportional Slowing 8 Patterns to 2 Patterns | The difference in mean time between presentation of the response stimulus options and the subject selecting the correct box on their first attempt on the 8 pattern assessment trials compared to the 2 pattern assessment trials. Calculated across 8 pattern and 2 pattern assessed trials where the first attempt was correct. | Complex (see description) | ms |  | 0 | ♾️ |
| MTS | MTSRCAMD | Reaction Time to Correct All Median | The median time between presentation of the response stimulus options and the subject selecting the correct box. Calculated across all assessed trials. | -ve | ms |  | 0 | ♾️ |
| MTS | MTSRFAMD | Reaction Time on First Attempt All Median | The median time between presentation of the response stimulus options and the subject selecting a box. Calculated across all assessed trials. | -ve | ms |  | 0 | ♾️ |
| OTS | **OTSMDLFC** | Median Latency to First Choice | The median latency, measured from the appearance of the stocking balls until the first box choice was made by the subject. Calculated across all assessed trials where the subject's first response was correct. | cx | ms |  | 0 | ♾️ |
| OTS | OTSPSFC | Problems Solved on First Choice | The total number of assessed trials where the subject chose the correct answer on their first attempt. Calculated across all assessed trials. | +ve | - |  | 0 | 15 |
| RVP | RVPA | RVP 'A | A’ (A prime) is the signal detection measure of a subject's sensitivity to the target sequence (string of three numbers), regardless of response tendency (the expected range is 0.00 to 1.00; bad to good). In essence, this metric is a measure of how good the subject is at detecting target sequences. | +ve | - |  | 0 | 1 |
| RVP | **RVPMDL** | RVP Median Response Latency | The median response latency on trials where the subject responded correctly. Calculated across all assessed trials. | -ve | ms |  | 100 | 1900 |
| RVP | RVPPFA | RVP Probability of False Alarm | The number of sequence presentations that were false alarms divided by the number of sequence presentations that were false alarms plus the number of sequence presentations that were correct rejections (False Alarms ÷ (False Alarms + Correct Rejections)) | -ve | - |  | 0 | 1 |
| SWM | SWMBE | SWM Between Errors | The number of times the subject incorrectly revisits a box in which a token has previously been found. Calculated across all assessed four, six and eight token trials. | -ve  (depends, in part, on the stage of the task reached) | - |  | 0 | 63 |
| SWM | SWMS | SWM Strategy (6-8 Boxes) | The number of times a subject begins a new search pattern from the same box they started with previously. If they always begin a search from the same starting point, we infer that the subject is employing a planned strategy for finding the tokens. Therefore, a low score indicates high strategy use (1 = they always begin the search from the same box), a high score indicates that they are beginning their searches from many different boxes. Calculated across assessed trials with 6 tokens or more. | -ve | - |  | 2 | 14 |

Supplenmentary Table 3. Results of multiple regression analysis with the CANTAB markers as dependent variables and the acute phase protein (APP) index, adverse childhood experiences (ACEs), metabolic and demographic variables as predictors.

|  |  | Coefficient statistics | | | Model statistics | | | |
| --- | --- | --- | --- | --- | --- | --- | --- | --- |
| Dependent variables | Explanatory variables | beta | t | p | R2 | F | df | p |
| DMS_PC0 | Age (years) | -0.344 | -4.069 | <.001 | 0.119 | 16.553 | 1, 124 | <.001 |
| ERT_UHRD | Age (years) | -0.403 | -5.04 | <.001 | 0.253 | 13.671 | 1, 124 | <.001^d^ |
|  | MetS ranking | -0.192 | -2.391 | 0.018 |  |  |  |  |
|  | APP index | -0.159 | -1.992 | 0.049 |  |  |  |  |
| OTS_MDLFC | Age (years) | 0.427 | 5.239 | <.001 | 0.182 | 27.451 | 1, 124 | <.001^b^ |
| RVP_MDL | Age (years) | 0.274 | 3.18 | 0.002 | 0.242 | 12.891 | 3, 124 | <.001^d^ |
|  | Education (years) | -0.236 | -2.761 | 0.007 |  |  |  |  |
|  | MetS | 0.18 | 2.226 | 0.028 |  |  |  |  |
| APP index | ACEs | 0.299 | 3.474 | <.001 | 0.089 | 12.071 | 1, 124 | <.001^b^ |
| MTS_PS82 | Age (years) | 0.481 | 6.085 | <.001 | 0.231 | 37.033 | 1, 124 | <.001^b^ |
| MTS_FAMD | Age (years) | 0.483 | 6.289 | <.001 | 0.397 | 19.791 | 4, 124 | <.001^e^ |
|  | MetS ranking | 0.166 | 2.267 | 0.025 |  |  |  |  |
|  | Education (years) | -0.182 | -2.343 | 0.021 |  |  |  |  |
|  | Transferiin | 0.151 | 2.077 | 0.04 |  |  |  |  |
| ERT_UHRS | Age (years) | -0.561 | -7.52 | <.001 | 0.315 | 56.556 | 1, 124 | <.001^b^ |
| CGT_DMQMT | WC (cm) | -0.237 | -2.682 | 0.008 | 0.077 | 5.116 | 2, 124 | .007^c^ |
|  | ACEs | -0.193 | -2.183 | 0.031 |  |  |  |  |

DMS_PC0, percent correct (0 second delay) of Delayed Match to Sample test; ERT_UHRD, unbiased hit rate disgust of Emotion Recognition Task; OTS_MDLFC, median latency to first choice of One Touch Stocking of Cambridge task; RVP_MDL, median response latency of Rapid Visual Processing task; APP index, acute phase protein index; ACEs, adverse childhood experiences; MTS_PS82, proportional slowing 8 patterns to 2 patterns of Match to Sample task; CGT_DMQMT, decisin making quality total merged of Cambridge Gambling Task.

Supplenmentary Table 4. Clinical scores in patients with major depressive disorder (MDD) and healthy controls (HCs).

|  | HC | | MDD | |
| --- | --- | --- | --- | --- |
|  | Mean | SD | Mean | SD |
| OSOD | -1.381 | 0.302 | 0.507 | 0.610 |
| Affective phenome | -1.403 | 0.272 | 0.598 | 0.603 |
| Physio-somatic phenome | -1.295 | 0.419 | 0.475 | 0.676 |
| Vegetative phenome | -1.378 | 0.353 | 0.591 | 0.588 |
| Current SI | -0.795 | 0.249 | 0.276 | 1.015 |
| Melancholia | -1.251 | 0.399 | 0.435 | 0.746 |
| Neuroticism | -1.077 | 0.612 | 0.392 | 0.807 |
| ROI | -0.885 | 0.525 | 0.308 | 0.942 |

OSOD, overall severity of depression; SI, suicidal ideation; ROI, recurrence of illness.
